# Glucagon-like peptide-1 receptor agonists and healthcare use in older adults with heart failure and obesity

**DOI:** 10.64898/2026.07.24.26358897

**Authors:** Clement Acheampong, Andy Bowe, Alexandra Hames, Monica Diaz, Mary Hayes, Insiya Poonawalla

## Abstract

**Background:** Glucagon-like peptide 1 receptor agonists (GLP-1 RA) are indicated for weight reduction and offer cardiometabolic health benefits, yet there is little real-world evidence regarding their association with healthcare resource utilization (HCRU) and costs for Medicare Advantage (MA) beneficiaries with heart failure and obesity.

**Methods:** We used a prevalent new-user study design and the Humana Healthcare Research database to identify MA beneficiaries with heart failure (HF) and obesity receiving standard HF therapy or HF therapy + newly initiating GLP-1 RA between January 1, 2022, and December 31, 2023. We observed adjusted, one-year mean difference in all-cause and heart failure-related HCRU.

**Results:** In our cohort of 4,677 matched pairs (mean age 72 years, male: 41%), one-year, adjusted risk ratio (RR) for HF therapy + GLP-1 RA vs HF therapy was 0.89 (95% CI: 0.84 – 0.93) for all-cause inpatient utilization, 0.88 (95% CI: 0.82 – 0.94) for avoidable hospitalizations, and 0.99 (95% CI: 0.95 – 1.03) for emergency department visits. For individuals receiving HF therapy + GLP-1 RA vs HF therapy, the mean difference was 30.6% (95% CI: 26.7% –34.7%) for all-cause total costs, 105.9% (95% CI: 99.1% –112.9%) for pharmacy costs, and −4.7% (95% CI: −8.7% to −0.6%) for medical costs. HF-related HCRU measures were lower among beneficiaries augmenting HF therapy with GLP-1 RA vs individuals receiving HF therapy alone.

**Conclusions:** Among MA beneficiaries with heart failure and obesity, the addition of GLP-1 RA to standard heart failure therapy reduced the likelihood of inpatient admissions or avoidable hospitalizations compared with HF therapy alone.

**Clinical perspective:** *What is new?:*

- Among older Medicare beneficiaries with heart failure and obesity, augmenting standard HF therapy with GLP-1 RA was associated with better disease control, less need for acute care, and lower medical costs, but increased all-cause expenditure due to higher medication costs.
- This study builds upon prior analyses by examining newer GLP-1 RA agents, including semaglutide and tirzepatide, and an older adult population at higher risk for poor outcomes.

*What are the clinical implications?:*

- This real-world evidence study suggests that the health benefits of GLP-1 RA use, as demonstrated by clinical trials, extend to observed improvements in utilization and economic outcomes.

## Introduction

Heart failure (HF) places an enormous burden on healthcare resources in the United States. Approximately 6.7 million Americans aged >20 years have HF, which is projected to reach 11.4 million by 2050.^1^ Annual HF-related U.S. expenditures are expected to rise from $30.7 billion in 2012 to $70 billion by 2030.^2^ HF also is associated with substantial morbidity and mortality and is a leading cause of hospitalization among Medicare beneficiaries.^3^ Obesity, a modifiable risk factor for HF development and progression,^4^ is prevalent in about 39% of adults ≥60 years.^5^ This condition is present in approximately 80% of patients with HF with preserved ejection fraction (HFpEF), where it exacerbates disease progression and significantly impacts healthcare use.^6^ The frequent co-occurrence of HF and obesity, particularly in older adults, drives substantial patient, clinical, and economic burden.^7^

For treatment of HF with reduced ejection fraction (HFrEF), guideline-directed medical therapy (GDMT) of renin–angiotensin system inhibitors or angiotensin receptor-neprilysin inhibitors, beta-blockers, mineralocorticoid receptor antagonists, and sodium-glucose cotransporter-2 inhibitors (SGLT-2i) is recommended to reduce morbidity and mortality^3,8^ For patients with HFpEF, treatment options have historically been limited; however, SGLT-2i provides clinical benefit in reducing hospitalizations and improving quality of life.^9^

Glucagon-like peptide-1 receptor agonists (GLP-1 RA) provide glycemic control, weight-reduction, and cardiometabolic benefits,^10^ with recent evidence suggesting cardiovascular benefits, particularly for individuals with HFpEF and obesity.^11,12^ The STEP-HFpEF trial demonstrated the GLP-1 RA semaglutide improves symptoms and exercise function while reducing physical limitations and induces greater weight loss than placebo in patients with HFpEF and obesity.^11^ Similar benefits were observed among patients with HFpEF, obesity, and type 2 diabetes.^12^

Despite demonstrated efficacy in clinical trials, real-world evidence on the impact of GLP-1 RA on healthcare resource utilization (HCRU) and costs in Medicare Advantage beneficiaries with HF and obesity remains limited. A recent real-world study of a younger, commercially insured population showed patients with HF and obesity/overweight who initiated semaglutide had 22% lower total medical costs at one-year follow-up ($6,512 difference, *P*=.001).^13^ However, their results may not be generalizable to older adults with multiple comorbidities. There is evidence tirzepatide offers similar clinical benefits to semaglutide among patients with HFpEF,^14^ but real world evidence examining HCRU and costs is limited to semaglutide.

In this study, we compared HCRU and costs in Medicare Advantage beneficiaries augmenting standard heart failure therapy with GLP-1 RA versus individuals maintaining standard heart failure therapy. This study examines real-world evidence for GLP-1 RA on managing HF and obesity in older adults.

## Methods

### Study Design, Data Source and Study Population

We used a prevalent new-user (PNU) design to investigate the effect of adding GLP-1 RA to standard HF medications on all-cause and heart failure HCRU and costs, compared with standard HF medications only.^15^ This design emulates a randomized controlled trial, in which participants on standard HF therapy are randomized to either continue the standard HF medications alone or augment therapy with a GLP-1 RA.

We used data from 2021 to 2024 from the Humana Healthcare Research Database, which includes demographic information, medical claims, and pharmacy claims for beneficiaries in Medicare Advantage with Prescription Drug (MAPD) plans (eMethods). This study did not constitute human subjects research and did not require institutional review board oversight, as determined by the Humana Human Subject Protection Office, which uses HHS regulations 45 CFR 46 and the Office for Human Research Protections guidance on Coded Private Information or Specimens Use in Research, Guidance (2008). This study followed the Strengthening the Reporting of Observational Studies in Epidemiology (STROBE) reporting guideline for cohort studies.^16^

The base cohort for this study included all MAPD plan beneficiaries aged 65-87 years who had one inpatient or two outpatient claims with *International Classification of Diseases, Tenth Revision, Clinical Modification (ICD-10-CM)* diagnosis codes for HF between an identification period of January 1, 2022, and December 31, 2023 (Figure 1). The date of the first claim was defined as the base cohort entry date.

**Figure 1.**
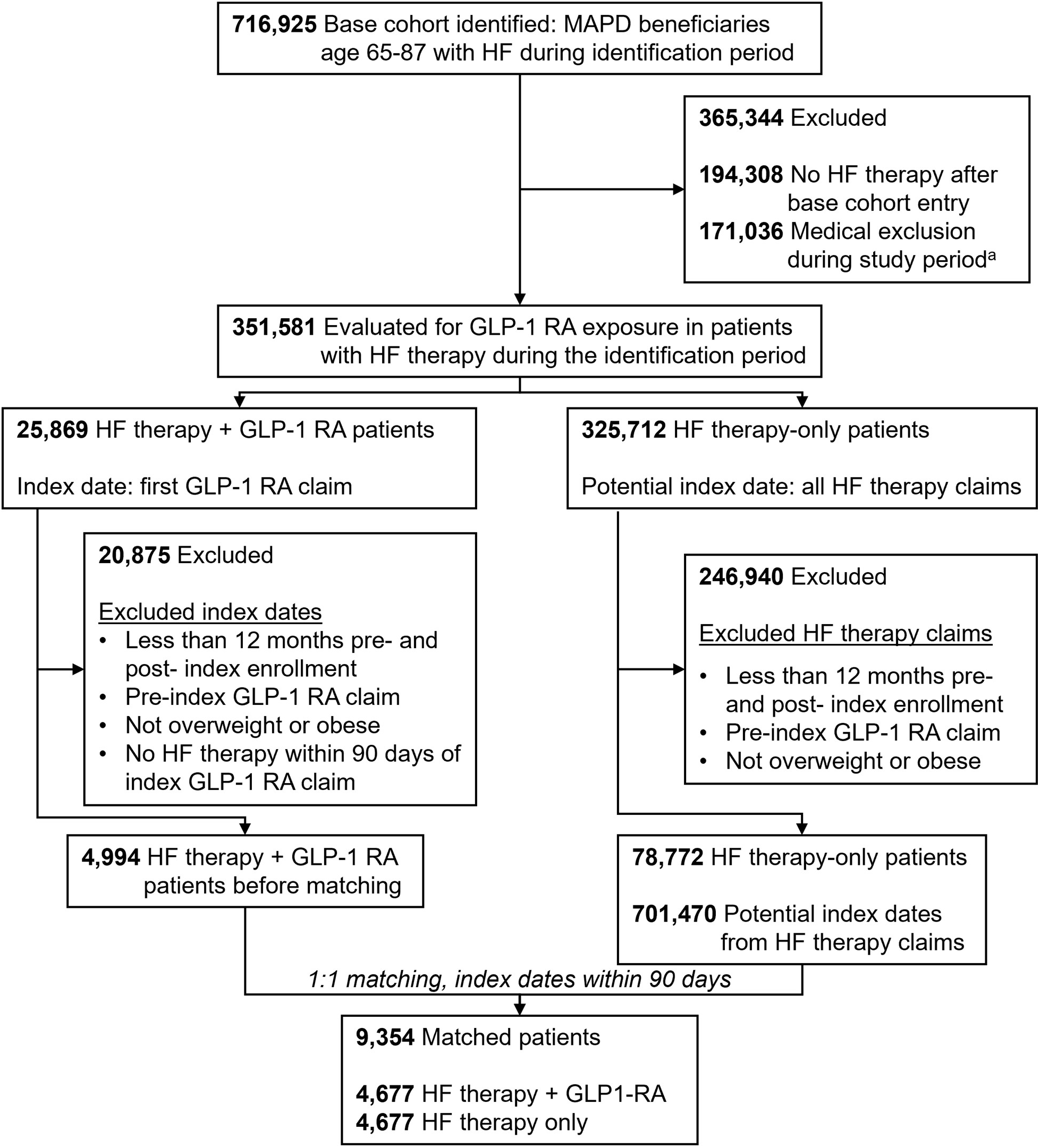
Attrition of Study Population. MAPD=Medicare Advantage Prescription Drug Plan; HF=Heart failure; GLP-1 RA= Glucagon-like peptide 1 receptor agonists ^a^Medical exclusions: patients with ICD-10-CM diagnosis codes on medical claims during the study period for Type 1 diabetes *(E10)*, pregnancy *(Z33)*, human immunodeficiency virus *(B20)*, heart transplant *(Z94.1)*, drug misuse *(F11-F19, T40, R78.1-R78.5),* chronic kidney disease *(I12.0, I13.11, I13.2, N18.5, N18.6, Z49.0, Z94.0, Z99.2),* end-stage illness *(E40-E43, F01-F03, G30, L89, R40.2, R64, Z59.3) and* nursing home resident *(Z59.3)*

### Exposure

We used an initial treatment (intention-to-treat) approach to define exposure groups, whereby patients were classified based on first treatment qualifying event, and subsequent treatment changes during follow-up were not considered.

#### HF therapy + GLP-1 RA treatment group

From the base cohort, we identified the treatment group as patients who newly initiated GLP-1 RA as an add-on to standard HF medications (HF therapy + GLP-1 RA) during the identification period, following the date patients first entered the base cohort (base cohort entry date). Patients were required to have at least two fills for GLP-1 RA therapies and evidence of concurrent standard HF medication within 90 days of GLP-1 RA initiation. The date of the first fill for a GLP-1 RA medication was designated as the index date. We excluded patients with a GLP-1 RA fill in the 12 months prior to index. Standard HF medication was operationally defined as use of a renin–angiotensin system inhibitors or angiotensin receptor-neprilysin inhibitors, beta-blockers, mineralocorticoid receptor antagonists, and/or SGLT-2 inhibitors, as mono, dual, triple or quadruple therapy, identified using Generic Product Identifier drug codes.

#### HF therapy-only group

For each person initiating GLP-1 RA, we constructed a set of potential control patients from the base cohort (HF therapy-only group), consisting of patients who had a fill for standard HF therapy within ±90 days of the index date for the corresponding patient taking HF therapy + initiating a GLP-1 RA and who had not filled a GLP-1 RA medication during the identification period. The corresponding standard HF therapy fill date was considered the potential index date for control patients.

All treatment and potential control patients were required to have continuous enrollment in MAPD for 12 months pre- and post-index and ICD-10-CM diagnosis codes for obesity in the 12-months pre-index (eTable 1). Patients with ICD-10-CM diagnosis codes for Type 1 diabetes, pregnancy, human immunodeficiency virus, heart transplant, drug misuse, chronic kidney disease stage V, end stage renal disease or kidney transplant, nursing home residence, or terminal end-stage illnesses were excluded (Figure 1).

### Outcomes

We assessed all-cause and HF-related healthcare resource utilization and costs over 12-months post-index. Inpatient care episodes were derived from facility claims using bill type, revenue codes, and dates of service. Emergency department (ED) visits were identified using revenue codes, place of treatment codes, and Current Procedural Terminology/Healthcare Common Procedure Coding System codes. HF-related visits were defined by the presence of an ICD-10-CM diagnosis code of I50 in the primary, second, or third position in the claim. Avoidable hospitalization was measured using the Prevention Quality Indicator from the Agency of Healthcare Research and Quality.^17^ Medical, pharmacy, and total costs were defined based on health plan total allowed amount and beneficiary paid costs and were adjusted to 2024 dollars using the Medical Consumer Price Index.

### Covariates

Covariates were measured in the 12 months pre-index. The covariates assessed included age, sex, race and ethnicity, geographic location, population density^18^, Medicare-Medicaid dual enrollment status, health plan type, comorbid conditions (eg, atherosclerotic cardiovascular disease, chronic obstructive pulmonary disease), overall comorbidity burden (Elixhauser Index, Rx-Risk-V score, Diabetes Complications Severity Index),^19–22^ body mass index, medication use, metrics of healthcare utilization, and costs (eTable 1).

### Statistical analysis

We used propensity score matching to balance differences in baseline covariates (eTable 1) between treatment and control groups. Time-conditional propensity scores were estimated using a Cox regression model stratified by calendar month of GLP-1 RA initiation. These scores were the cumulative probability of either adding a GLP-1 RA or continuing the standard HF medications following cohort entry for each patient. Patients were matched 1:1 with replacement, using the greedy caliper matching method, with a caliper width equal to 0.2 of the SD of the logit of the propensity score.^23^ Baseline population demographics, clinical characteristics, and medication use for the unmatched and matched comparison cohorts were described using means (SD) for continuous measures and frequencies (%) for categorical variables. We evaluated the adequacy of covariate balance after matching based on standardized mean differences (SMD), with a threshold of less than 0.1 indicating satisfactory balance.^24^

HCRU outcomes were compared between the HF + GLP-1 RA treatment group and the matched HF therapy-only controls using matched-pair tests (McNemar’s paired test for binary measures and Wilcoxon signed rank test for continuous measures) and reported the adjusted relative risk ratio with corresponding 95% confidence intervals. We modeled costs using generalized linear regression and a γ distribution with a log-link function to account for skewness in healthcare spending distributions. Pre-index cost measures were included in all regression models. We reported the exponentiated estimates and the mean percentage differences with corresponding 95% CIs.^25^ All analyses were conducted using SAS Enterprise Guide 8.3.

### Sensitivity analyses

To examine the robustness of our primary results, we assessed outcomes among patients without a HF diagnosis code (ICD-10-CM code I50) in the 12-month pre-index period to proxy newly diagnosed HF cases.

## Results

A total of 4,994 patients with HF therapy + GLP-1 RA and 701,470 potential controls were eligible for study inclusion; after applying propensity score matching, both study groups included 4,677 patients (Table 1). Before matching, the HF therapy + GLP-1 RA were younger (mean age 71.99 vs. 74.91), predominantly female (58.99% vs. 52.71%), and had a lower proportion of white patients (68.02% vs. 71.33%) compared with the HF therapy-only group. Additionally, patients on HF therapy who initiated a GLP-1 RA were less likely to have atrial fibrillation (29.62% vs. 38.48%) and more likely to have type 2 diabetes (91.29% vs. 55.57%), diabetes complications (nephropathy, 12.13% vs. 6.07%; neuropathy, 48.50% vs. 24.26%; retinopathy, 21.51% vs. 10.69%), and obstructive sleep apnea (46.94% vs. 29.58%). After matching, the exposure groups were well-balanced across all covariates, with SMD < 0.1, indicating successful adjustment for these potential confounders.

**Table 1.** Baseline Demographic and Clinical Characteristics, Unmatched and Matched Cohorts.

| Patient Characteristics | Before Matching<br>(All Eligible Control Events Included) |  | After Matching (1:1) |  | Standardized<br>Mean<br>Differences<br>(SMD) |
| --- | --- | --- | --- | --- | --- |
|  | HF therapy only<br>n=701,470 | HF therapy +<br>GLP-1 RA<br>n=4,994 | HF therapy only<br>n=4,677 | HF therapy +<br>GLP-1 RA<br>n=4,677 |  |
| **Age** |  |  |  |  |  |
| Mean (SD) | 74.91 (5.4) | 71.99 (4.69) | 72.12 (4.64) | 72.04 (4.70) | 0.017 |
| Median [IQR] | 75 [71-79] | 71 [68-75] | 71 [68-75] | 71 [68-75] |  |
| **Sex, n (%)** |  |  |  |  |  |
| Female | 369,738 (52.71%) | 2,946 (58.99%) | 2,697 (57.67%) | 2,744 (58.67%) | 0.020 |
| Male | 331,708 (47.29%) | 2,048 (41.01%) | 1,980 (42.33%) | 1,933 (41.33%) | 0.020 |
| **Geographic Region, n (%)** |  |  |  |  |  |
| Northeast | 23,044 (3.29%) | 146 (2.92%) | 115 (2.46%) | 134 (2.87%) | 0.025 |
| Midwest | 114,887 (16.38%) | 875 (17.52%) | 797 (17.04%) | 808 (17.28%) | 0.006 |
| South | 508,957 (72.56%) | 3,540 (70.89%) | 3,350 (71.63%) | 3,330 (71.20%) | 0.009 |
| West | 54,558 (7.78%) | 433 (8.67%) | 415 (8.87%) | 405 (8.66%) | 0.008 |
| **Population Density, n (%)** |  |  |  |  |  |
| Rural | 47,991 (6.84%) | 373 (7.47%) | 356 (7.61%) | 347 (7.42%) | 0.007 |
| Non-Rural | 653,479 (93.16%) | 4,621 (92.53%) | 4,321 (92.39%) | 4,330 (92.58%) | 0.007 |
| **Race, n (%)** |  |  |  |  |  |
| White | 500,385 (71.33%) | 3397 (68.02%) | 3,204 (68.51%) | 3,183 (68.06%) | 0.010 |
| Black | 156,343 (22.29%) | 1,277 (25.57%) | 1,175 (25.12%) | 1,194 (25.53%) | 0.009 |
| Underrepresented Race/Ethnicity<sup>a</sup> | 34,519 (4.92%) | 254 (5.09%) | 233 (4.98%) | 237 (5.07%) | 0.004 |
| Unknown<sup>b</sup> | 10,223 (1.46%) | 66 (1.32%) | 65 (1.39%) | 63 (1.35%) | 0.004 |
| **Low Income Subsidy/Dual Eligibility Status, n (%)** |  |  |  |  |  |
| No LIS or DE Status | 210,941 (30.07%) | 2,012 (40.29%) | 1,820 (38.91%) | 1,860 (39.77%) | 0.018 |
| LIS status and/or Medicare-Medicaid DE | 490,529 (69.93%) | 2,982 (59.71%) | 2,857 (61.09%) | 2,817 (60.23%) | 0.018 |
| **Health Plan Type, n (%)** |  |  |  |  |  |
| HMO | 355,667 (50.72%) | 2,180 (43.65%) | 2,085 (44.58%) | 2,060 (44.05%) | 0.011 |
| PPO | 197,954 (28.23%) | 1,422 (28.47%) | 1,368 (29.25%) | 1,327 (28.37%) | 0.019 |
| Other (Including POS and FFS) | 33,896 (4.83%) | 1,392 (27.87%) | 1,224 (26.17%) | 1,290 (27.58%) | 0.032 |
| **Year of Index Date, n (%)** |  |  |  |  |  |
| 2022 | 269,013 (38.35%) | 1,394 (27.91%) | 1,458 (31.17%) | 1,379 (29.48%) | 0.037 |
| 2023 | 432,457 (61.65%) | 3,600 (72.09%) | 3,219 (68.83%) | 3,298 (70.52%) | 0.037 |
| **Number of Days Between Cohort Entry and Index Date** |  |  |  |  |  |
| Mean (SD) | 294.24 (194.2) | 300.7 (189.8) | 293.97 (187.38) | 293.97 (187.38) | 0.000 |
| Median [IQR] | 275 [125-449] | 297.5 [135-450] | 287 [133-439] | 287 [133-439] |  |
| **Elixhauser Comorbidity Index** |  |  |  |  |  |
| Mean (SD) | 5.95 (2.43) | 6.18 (2.29) | 6.29 (2.29) | 6.28 (2.26) | 0.007 |
| Median [IQR] | 6 [4-7] | 6 [5-8] | 6 [5-8] | 6 [5-8] |  |
| **RxRisk-V Score** |  |  |  |  |  |
| Mean (SD) | 8.01 (2.77) | 8.87 (2.78) | 8.90 (2.81) | 8.88 (2.78) | 0.009 |
| Median [IQR] | 8 [6-10] | 9 [7-11] | 9 [7-11] | 9 [7-11] |  |
| **Diabetes Complications Severity Index** |  |  |  |  |  |
| Mean (SD) | 2.94 (1.49) | 3.19 (1.64) | 3.29 (1.59) | 3.29 (1.61) | 0.000 |
| Median [IQR] | 3 [2-4] | 3 [2-4] | 3 [2-4] | 3 [2-4] |  |
| **Baseline BMI (Closest Pre-Index)** |  |  |  |  |  |
| Overweight (BMI 25 - <30) | 167,611 (23.89%) | 361 (7.23%) | 328 (7.01%) | 344 (7.36%) | 0.013 |
| Obesity Class 1 (BMI 30 - <35) | 168,951 (24.09%) | 876 (17.54%) | 866 (18.52%) | 835 (17.85%) | 0.017 |
| Obesity Class 2 (BMI 35 - <40) | 112,222 (16%) | 1,039 (20.8%) | 961 (20.55%) | 984 (21.04%) | 0.012 |
| Obesity Class 3 (BMI 40+) | 118,062 (16.83%) | 1,880 (37.65%) | 1,744 (37.29%) | 1,754 (37.50%) | 0.004 |
| Other or Unspecified Obesity | 134,624 (19.19%) | 838 (16.78%) | 778 (16.63%) | 760 (16.25%) | 0.010 |
| **Baseline ASCVD Categories, n (%)** |  |  |  |  |  |
| Any Pre-Index ASCVD Events | 595,219 (84.85%) | 4,179 (83.68%) | 3,953 (84.52%) | 3,948 (84.41%) | 0.003 |
| Established ASCVD: 1+ IP or 2+ OP Days | 524,339 (74.75%) | 3,634 (72.77%) | 3,452 (73.81%) | 3,447 (73.70%) | 0.002 |
| MI/ACS | 134,890 (19.23%) | 793 (15.88%) | 735 (15.72%) | 755 (16.14%) | 0.012 |
| Carotid Artery Stenosis | 89,931 (12.82%) | 569 (11.39%) | 551 (11.78%) | 549 (11.74%) | 0.001 |
| Coronary Artery Disease | 398,385 (56.79%) | 2,725 (54.57%) | 2,572 (54.99%) | 2,581 (55.18%) | 0.004 |
| Angina | 188,812 (26.92%) | 1,374 (27.51%) | 1,317 (28.16%) | 1,310 (28.01%) | 0.003 |
| Stroke | 55,098 (7.85%) | 326 (6.53%) | 330 (7.06%) | 318 (6.80%) | 0.010 |
| Transient Ischemic Attack | 68,518 (9.77%) | 424 (8.49%) | 421 (9.00%) | 411 (8.79%) | 0.008 |
| Aortic Artherosclerotic Disease | 218,675 (31.17%) | 1,269 (25.41%) | 1,252 (26.77%) | 1,222 (26.13%) | 0.015 |
| Peripheral Arterial Disease | 324,934 (46.32%) | 2,539 (50.84%) | 2,373 (50.74%) | 2,414 (51.61%) | 0.018 |
| Revascularization (Coronary and Arterial) | 29,851 (4.26%) | 182 (3.64%) | 169 (3.61%) | 176 (3.76%) | 0.008 |
| **Baseline Comorbidities, n (%)** |  |  |  |  |  |
| Lower Extremity Amputation | 6,843 (0.98%) | 78 (1.56%) | 80 (1.71%) | 76 (1.62%) | 0.007 |
| Atrial fibrillation | 269,928 (38.48%) | 1,479 (29.62%) | 1,431 (30.60%) | 1,430 (30.58%) | 0.000 |
| Type 2 diabetes | 389,778 (55.57%) | 4,559 (91.29%) | 4,334 (92.67%) | 4,269 (91.28%) | 0.051 |
| Nephropathy | 42,588 (6.07%) | 606 (12.13%) | 541 (11.57%) | 578 (12.36%) | 0.024 |
| Neuropathy | 170,190 (24.26%) | 2,422 (48.5%) | 2,288 (48.92%) | 2,289 (48.94%) | 0.000 |
| Retinopathy | 74,955 (10.69%) | 1,074 (21.51%) | 994 (21.25%) | 1,005 (21.49%) | 0.006 |
| Chronic kidney disease | 324,466 (46.26%) | 2,307 (46.2%) | 2,173 (46.46%) | 2,216 (47.38%) | 0.018 |
| NASH/NAFLD | 39,837 (5.68%) | 454 (9.09%) | 394 (8.42%) | 415 (8.87%) | 0.016 |
| Obstructive sleep apnea | 207,523 (29.58%) | 2,344 (46.94%) | 2,173 (46.46%) | 2,196 (46.95%) | 0.010 |
| Chronic obstructive pulmonary disease | 222,363 (31.7%) | 1,729 (34.62%) | 1,667 (35.64%) | 1,636 (34.98%) | 0.014 |
| Depression | 104,698 (14.93%) | 971 (19.44%) | 945 (20.21%) | 909 (19.44%) | 0.019 |
| Pancreatic disorders | 14,529 (2.07%) | 99 (1.98%) | 84 (1.80%) | 89 (1.90%) | 0.008 |
| **Baseline Heart Failure Medications, n (%)** |  |  |  |  |  |
| ACEi | 252,098 (35.94%) | 1,680 (33.64%) | 1,542 (32.97%) | 1,565 (33.46%) | 0.010 |
| ARB | 316,818 (45.16%) | 2,356 (47.18%) | 2,250 (48.11%) | 2,205 (47.15%) | 0.019 |
| ARNi | 88,894 (12.67%) | 384 (7.69%) | 384 (8.21%) | 382 (8.17%) | 0.002 |
| BB | 471,828 (67.26%) | 2,815 (56.37%) | 2,626 (56.15%) | 2,689 (57.49%) | 0.027 |
| MRA | 177,540 (25.31%) | 1,035 (20.72%) | 998 (21.34%) | 1,001 (21.40%) | 0.002 |
| SGLT2i | 106,385 (15.17%) | 1,264 (25.31%) | 1,206 (25.79%) | 1,193 (25.51%) | 0.006 |
| Loop Diuretics | 393,049 (56.03%) | 3,080 (61.67%) | 2,862 (61.19%) | 2,921 (62.45%) | 0.026 |
| **Heart Failure Therapy, n (%)** |  |  |  |  |  |
| None | 7,022 (1.00%) | 78 (1.56%) | 74 (1.58%) | 69 (1.48%) | 0.009 |
| Monotherapy | 225,329 (32.12%) | 1,878 (37.61%) | 1,685 (36.03%) | 1,718 (36.73%) | 0.015 |
| Dual Therapy | 304,095 (43.35%) | 1,952 (39.09%) | 1,890 (40.41%) | 1,836 (39.26%) | 0.024 |
| Triple Therapy | 130,352 (18.58%) | 850 (17.02%) | 822 (17.58%) | 822 (17.58%) | 0.000 |
| Quad Therapy | 34,672 (4.94%) | 236 (4.73%) | 206 (4.40%) | 232 (4.96%) | 0.026 |

HF= Heart Failure; GLP-1 RA= Glucagon-like peptide 1 receptor agonists; LIS= low income subsidy; DE=dual-eligible, HMO=health maintenance organization, PPO=preferred provider organization, POS=point of sale, FFS=fee-for-service, SD=standard deviation, IQR=interquartile range, ASCVD = Atherosclerotic Cardiovascular Disease, NASH = Nonalcoholic Steatohepatitis, NAFLD = Nonalcoholic Fatty Liver Disease, ACEi = Angiotensin-converting enzyme inhibitors, ARB = Angiotensin II Receptor Blockers, ARNi = Angiotensin receptor-neprilysin inhibitors, BB = Beta-adrenergic blocking agents, MRA = Mineralocorticoid receptor antagonists, SGLT2i = Sodium-glucose transport 2 inhibitors
<sup>a</sup>Includes Hispanic, Asian, American Indian or Alaska Native, Pacific Islander, or Other
<sup>b</sup>Missing race or ethnicity indicator data in the CMS administrative database
Cohorts adjusted for baseline differences in index date and demographic, clinical and utilization characteristics with propensity score matching. Balance between matched cohorts was assessed using standardized mean differences (SMD); all differences in baseline demographic and clinical characteristics had calculated SMD<0.1 indicating the cohorts were well-balanced.

Over the 12-month follow-up period, patients on HF therapy + GLP-1 RA had reduced incidence of all-cause inpatient utilization compared with patients continuing on HF therapy alone (18.30% vs. 22.22%; *P* < 0.001, Table 2), corresponding to an adjusted risk ratio (RR) of 0.89 (95% CI: 0.85 – 0.93, Figure 2). Avoidable hospitalizations occurred less frequently among patients initiating GLP-1 RAs (6.41% vs. 8.30%; *P* = 0.001, Table 2), with an RR of 0.88 (95% CI: 0.82 – 0.94, Figure 2). The proportion of patients with at least one all-cause ED visit was comparable between groups (44.43% vs. 44.96%; *P* = 0.602; RR = 0.99, 95% CI: 0.95 – 1.03). Patterns were similar when evaluating HF-specific HCRU (Table 2). HF-related inpatient visits were lower among individuals on HF therapy and initiating GLP-1 RA compared with patients continuing HF therapy alone (6.78% vs. 9.17%; *P* < 0.001), with an associated RR of 0.86 (95% CI: 0.80 – 0.92). HF-related ED visits were lower in the HF therapy + GLP-1 RA group (9.60% vs. 11.95%; *P* < 0.001), corresponding to an RR of 0.89 (95% CI: 0.84 – 0.94).

**Figure 2.**
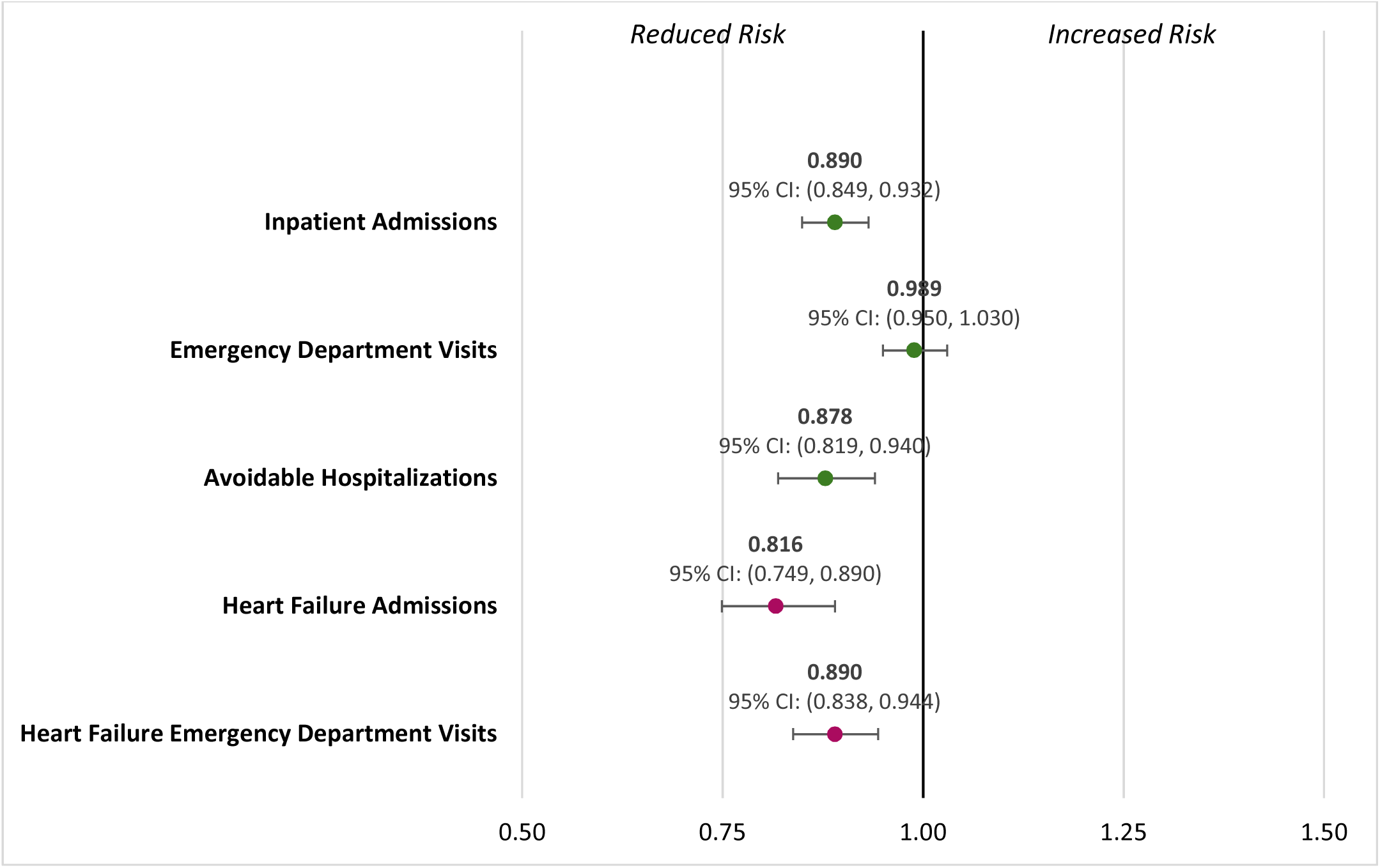
Risk Ratios for Healthcare Resource Utilization in 12 months Post-Index Period.

**Table 2.** All-Cause and Heart Failure-Related Healthcare Resource Utilization Outcomes in 12-Month Post-Index Period.

| **Patient Outcomes** | **HF therapy only**<br>**n=4,677** | **After Matching (1:1)**<br>**HF therapy + GLP-1 RA**<br>**n=4,677** | **P Value** |
| --- | --- | --- | --- |
| **Inpatient Hospitalization** |  |  |  |
| n (%) | 1,039 (22.22%) | 856 (18.30%) | **<0.001** |
| Mean (SD) | 1.48 (0.94) | 1.39 (0.77) | **<0.001** |
| **Emergency Department Visit** |  |  |  |
| n (%) | 2,103 (44.96%) | 2,078 (44.43%) | 0.602 |
| Mean (SD) | 2.14 (1.81) | 2.04 (1.93) | 0.08 |
| **Avoidable Hospitalization** |  |  |  |
| n (%) | 388 (8.30%) | 300 (6.41%) | **0.001** |
| Mean (SD) | 1.37 (0.73) | 1.36 (0.72) | **0.001** |
| **Heart Failure Admissions** |  |  |  |
| n (%) | 215 (4.60%) | 139 (2.97%) | **<0.001** |
| Mean (SD) | 1.34 (0.66) | 1.43 (0.83) | **0.001** |
| **Heart Failure Emergency Department Visit** |  |  |  |
| n (%) | 559 (11.95%) | 449 (9.60%) | **<0.001** |
| Mean (SD) | 1.46 (0.87) | 1.4 (0.82) | **<0.001** |

HF= Heart Failure; GLP-1 RA= Glucagon-like peptide 1 receptor agonists; SD=standard deviation

During the 12-month follow-up period, average per-person per-month (PPPM) all-cause total costs were higher among the HF therapy + GLP-1 RA group compared with HF therapy alone ($2,878 vs. $2,289), representing a 30.6% adjusted mean difference between groups (95% CI: 26.7%–34.7%, Table 3). This difference was primarily driven by higher pharmacy costs. Specifically, the average mean PPPM pharmacy cost was $1,569 in the HF therapy + GLP-1 RA group compared with $900 among the HF therapy-only group, corresponding to a 105.9% adjusted mean difference (95% CI: 99.1%–112.9%). Average mean PPPM medical costs were modestly lower among patients in the HF therapy + GLP-1 RA initiation group ($1,310) than among patients continuing HF therapy alone ($1,390), yielding a −4.7% adjusted mean difference (95% CI: −8.7% to −0.6%). The HF therapy + GLP-1 RA group had lower average HF-related total ($363 versus $445 PPPM, −20.4% adjusted mean difference), pharmacy ($171 vs. $195 PPPM, −13.4% adjusted mean difference), and medical costs ($192 vs. $250 PPPM, −22.3% adjusted mean difference, Table 3).

**Table 3.**
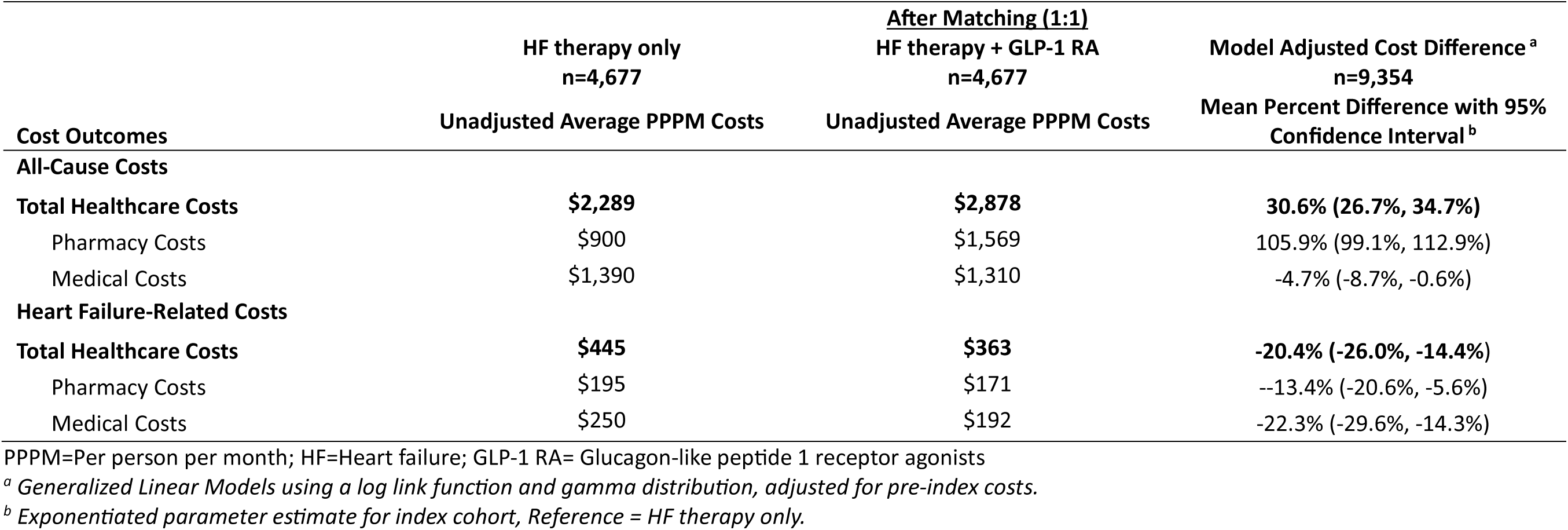
All-Cause and Heart Failure-Related Cost Outcomes in 12-Month Post-Index Period.

| Cost Outcomes | HF therapy only<br>n=4,677 | After Matching (1:1)<br>HF therapy + GLP-1 RA<br>n=4,677 | Model Adjusted Cost Difference <sup>a</sup><br>n=9,354 |
| --- | --- | --- | --- |
|  | Unadjusted Average PPPM Costs | Unadjusted Average PPPM Costs | Mean Percent Difference with 95%<br>Confidence Interval <sup>b</sup> |
| **All-Cause Costs** |  |  |  |
| **Total Healthcare Costs** | **\$2,289** | **\$2,878** | **30.6% (26.7%, 34.7%)** |
| Pharmacy Costs | \$900 | \$1,569 | 105.9% (99.1%, 112.9%) |
| Medical Costs | \$1,390 | \$1,310 | -4.7% (-8.7%, -0.6%) |
| **Heart Failure-Related Costs** |  |  |  |
| **Total Healthcare Costs** | **\$445** | **\$363** | **-20.4% (-26.0%, -14.4%)** |
| Pharmacy Costs | \$195 | \$171 | --13.4% (-20.6%, -5.6%) |
| Medical Costs | \$250 | \$192 | -22.3% (-29.6%, -14.3%) |

PPPM=Per person per month; HF=Heart failure; GLP-1 RA= Glucagon-like peptide 1 receptor agonists
<sup>a</sup> Generalized Linear Models using a log link function and gamma distribution, adjusted for pre-index costs.
<sup>b</sup> Exponentiated parameter estimate for index cohort, Reference = HF therapy only.

### Sensitivity analysis

After restricting the analysis to patients without an HF diagnosis in the 12 months prior to cohort entry, results were consistent with the primary findings and similar patterns were observed for both healthcare resource utilization and healthcare costs (eTable 2 and eTable 3).

## Discussion

In a large population of Medicare Advantage beneficiaries with HF and obesity, GLP-1 RA augmentation to standard HF therapy was associated with fewer all-cause and HF-related hospitalizations; lower HF-related ED use; and lower HF-related total, medical, and pharmacy costs. All-cause total costs were higher, driven primarily by pharmacy spending compared with standard HF therapy. These real-world findings expand on trial evidence^11,12^ showing improved utilization and economic outcomes with GLP-1 RA augmentation for older adults with obesity-related HF. Our results build on previous cost analyses in younger, commercially insured populations^13^ by evaluating an older MA population with HF and greater comorbidity burden and included users of both semaglutide and tirzepatide.

Our observed reduction in HF-related admissions among individuals receiving GLP-1 RA therapy is consistent with clinical trial evidence demonstrating improvements in symptoms, physical limitations, and exercise capacity in obesity-related HFpEF, which may translate into reduced decompensation risk.^12^ The effects of GLP-1 RA therapy on HF-related outcomes may be largely mediated through weight reduction. Weight loss has been associated with improvements in diastolic function, decreased inflammation and better glycemic control, providing a biologically plausible mechanism through which GLP-1 RA therapy could lower HF-related HCRU.^3^ Furthermore, our findings support the emerging view of HFpEF with obesity as a modifiable phenotype, and GLP-1 RA therapy may represent an effective intervention to target this population and improve clinical outcomes.^26^ Our work aligns with evolving policy around HF care which emphasizes focused management guided by cardiometabolic phenotype.^26^

The disease-specific reductions in medical cost we observed with GLP-1 RA use for HF patients suggest a shift in spending from acute HF care to pharmacotherapy. This highlights the importance of considering disease-specific cost offsets in formulary decisions rather than pharmacy expenditures in isolation. However, our work also highlights the possible trade-offs between higher pharmacy costs due to prices of GLP-1 RAs and reduced HCRU in other areas, particularly among HF patients. The recent Medicare Part D benefit redesign, which capped beneficiary out-of-pocket spending beginning in 2025 and increased Part D plan liability, may further increase GLP-1 RA spending. For beneficiaries, the reduced cost-related barriers likely induce greater initiation, treatment persistence, and potential for improved downstream health outcomes. However, reduced costs associated with improved outcomes such as HF hospitalization occur downstream, while health plans continue to face near-term cost-pressures from high drug prices. CMS policy initiatives including the Medicare Drug Price Negotiation Program (reducing price of semaglutide in 2027 from $959 to $274 for a 30-day supply) and innovation center models that expand coverage aim to reduce costs and improve access. Together, these policies may better align incentives across beneficiaries, plans, and manufacturers to realize the full clinical and economic value of GLP-1 RA therapies. This dynamic is particularly relevant for Medicare Advantage plans, where HF admissions are a major cost driver. ^2^

Our findings are consistent with existing literature and provide a foundation for future studies to further refine the optimal therapeutic management of HF in individuals with obesity. Future research could stratify by HF phenotype, diabetes status, and obesity class, thereby identifying sub-populations most likely to benefit from the addition of a GLP-1 RA to standard HF therapy. Evaluation of longer-term outcomes would offer greater insight into cost trajectories and net budget impact, particularly given HF is a chronic condition requiring life-long management, and may demonstrate additional medical cost offsets accrue with sustained GLP-1 RA use. Examining measures of treatment persistence and adherence in future analyses will be important, as sustained exposure to GLP-1 RAs may be necessary to realize their full cardiometabolic and economic benefits. Incorporating longitudinal clinical data, such as ejection fraction, natriuretic peptides, and weight changes would enable a more granular understanding of the potential mechanisms through which GLP-1 RAs exert protection against higher-intensity HCRU among patients with HF and obesity.

## Limitations

This study has limitations related to the use of observational administrative claims data. Potential misclassification due to coding inaccuracies, missing data, and variability in clinical documentation practices may affect the reliability of important measures (eg, obesity). However, we applied validated claims-based definitions where available and required multiple diagnosis codes to confirm key clinical conditions. We cannot rule out the presence of residual confounding due to unmeasured variables such as frailty, HF severity, ejection-fraction subtype, and New York Heart Association class. However, the prevalent new-user design was chosen to minimize immortal time bias,^15^ and time-conditional propensity score matching allowed for measured clinical, utilization, and cost covariate adjustments in analysis. We did not directly observe HF phenotype of HFpEF vs HFrEF, which could have led to enrichment for the obesity-related HFpEF group. Our use of an on-treatment approach for analysis may have introduced selection bias, and we could not measure drug adherence or weight loss after augmentation of standard HF therapy with a GLP-1 RA. Our inclusion of only MA beneficiaries may limit generalizability to younger populations, commercially insured individuals, and other fee-for-service models.

## Conclusions

GLP-1 RA augmentation of HF standard of care in HF with obesity was associated with fewer HF hospital events and lower HF-specific costs, with higher total costs driven by pharmacy spending, indicating a shift from acute HF care toward pharmacologic management.

## Data Availability

De-identified data that support the results of this study may be made available from the research team with: (1) agreement to collaborate with the study team on all publications, (2) provision of external funding for administrative and investigator time necessary for this collaboration, (3) demonstration that the external investigative team is qualified and has documented evidence of training for human subjects protections, and (4) agreement to abide by the terms outlined in data use agreements between institutions. Request for proprietary data, such as pricing information, would be considered on a case-by-case basis and may not be approved.

## Acknowledgments, Sources of Funding, & Disclosures

## Acknowledgements

The authors thank Suzanne Dixon, MPH, MS, RDN for medical writing support in drafting the discussion section and for editorial review of the manuscript. The authors maintained full control over the content, interpretation, and conclusions presented in the manuscript and approved the final version for submission.

